# Risk factors for SARS-CoV-2 transmission in student residences: a case-ascertained study in Leuven, Belgium from October 2020 until May 2021

**DOI:** 10.1101/2022.03.23.22272836

**Authors:** Marte Vanbesien, Geert Molenberghs, Caspar Geenen, Jonathan Thibaut, Sarah Gorissen, Emmanuel André, Joren Raymenants

**Affiliations:** Faculty of medicine, KU Leuven, Belgium; Interuniversity Institute for Biostatistics and statistical Bioinformatics, Data Science Institute, Hasselt University, Belgium; Interuniversity Institute for Biostatistics and statistical Bioinformatics, KU Leuven, Belgium; Department of Microbiology, Immunology and Transplantation, KU Leuven, Herestraat 49, 3000 Leuven, Belgium; Department of laboratory medicine, University Hospitals Leuven, Herestraat 49, 3000 Leuven, Belgium

**Author notes:** **Corresponding author:** Joren Raymenants.

**Keywords:** COVID-19, SARS-CoV-2, transmission, risk factors, student residence, shared household, congregate setting

## Abstract

**Background:** Student residences are at high risk for rapid COVID-transmission due to crowding and frequent close contact.

**Aim:** We aimed to investigate the overall secondary attack rates (SAR) in student residences and to discern risk factors for higher transmission in order to improve the evidence base for screening efforts and preventive measures.

**Methods:** In this retrospective case-ascertained study, we analysed data from student residences screened in Leuven, Belgium between October 2020 and May 2021, following detection of a COVID-19 case in the residence. We investigated the impact on the SAR in the living units screened of delay-time until follow-up, shared use of kitchen or sanitary facilities, presence of an external infection source and occurrence of social gatherings attended by the index case.

**Results:** We included 200 residence units, representing 2326 screened residents, of which 68 units showed secondary transmission. The overall SAR was estimated at 0.0813 (95%CI 0.0705-0.0936). Both sharing sanitary facilities (p=0.04) and social gatherings attended by the index case (p=0.033) significantly impacted SAR, which increased from 3% to 13% when both risk factors were present compared to absent.

**Conclusions:** We identify risk factors which should be considered when selecting students for screening during an outbreak of COVID-19 in student residences to improve comprehensiveness and proportionality of testing. The identified risk factors improve the evidence base for preventive measures aimed at limiting social gatherings and improving ventilation of shared spaces in outbreak-prone settings. Lastly, they should be considered when designing student accommodation and other shared households.

## INTRODUCTION

The COVID-19 pandemic, caused by the RNA virus SARS-CoV-2 first detected in Wuhan, China in December 2019 has caused over 6.1 million reported deaths as of March 2022^1^.

Despite the buildup of natural and vaccine-based immunity, widespread community transmission of COVID-19 continues to put pressure on health systems. Non-pharmaceutical interventions including isolation of confirmed cases, tracing and quarantining of contacts, and testing of symptomatic and at-risk individuals, are likely to remain essential to mitigate the overall impact of the pandemic for the foreseeable future^2^.

Congregate settings, such as curative and residential care settings, correctional facilities, and student residences, are at risk of rapid COVID-19 transmission due to crowding and frequent close contact^3^. They are suggested to initiate outbreaks that spill over to other high-risk settings and the community^4^. Student residences hold additional risk to the community since the age cohort they belong to has a larger average number of social contacts than other cohorts^5^.

Despite these risks, there is a paucity of data on the range of secondary attack rates (SAR) one may find in student residences, which risk factors underpin the observed variation in transmission and how they compare to regular households^6–8^. Some studies suggest a higher risk of transmission if residents share living spaces^9^ or if residents do not adhere to prevention measures^8^. However, the sample size in these studies is too small to draw any reliable conclusions. Studies examining COVID-19 transmission in the household setting identified the number of household contacts, the nature of relationship between contacts, the age of contacts and the presence of symptoms as risk factors for higher rates of secondary transmission^10^, but even in the household setting, there is only limited data about the influence of behaviour- or infrastructure-related factors on the secondary infection probability. One study observed a trend towards higher SAR in households if members kissed, hugged, shared sanitary facilities or a bed, although these results were not found to be significant^11^. Furthermore, it is unclear whether these supposed risk factors in household settings are transferable to other high-risk settings such as student residences. This lack of evidence base limits the implementation of effective preventive measures and a comprehensive testing strategy which complements effectiveness with proportionality to achieve efficient transmission reduction.

To fill this knowledge gap, we conducted a retrospective case-ascertainment study and analyzed 165 instances in which a student residence was screened in the student town of Leuven, Belgium, following the detection of at least one case of COVID-19 in the residence. We quantified the SAR overall and its range and collected information related to the living arrangements and interactions within and outside of the screened student residences to test whether they were associated with a higher SAR. In a sensitivity analysis, we investigated whether restricting the analysis to units harboring the initial index case altered our conclusions.

## METHODS

### Setting and Design

This retrospective case-ascertained study was performed on data gathered in the context of a testing and contact tracing system targeted to over 30.000 tertiary education students in Leuven, Belgium^12,13^. A standard screening protocol for student residences was introduced on October 30^th^ 2020 and aimed to strike a balance between effectiveness and proportionality (figure 1). All data from cases and contacts gathered during the follow-up of a residence outbreak were coded into a customized version of Go.Data. The inclusion and exclusion criteria for residences, residence units and individuals are described in Figure 2. A student residence was defined as an architectural complex housing mostly tertiary education students. A residence unit was defined as a group of student rooms within a residence that shared either sanitary or kitchen facilities.

**Figure 1:**
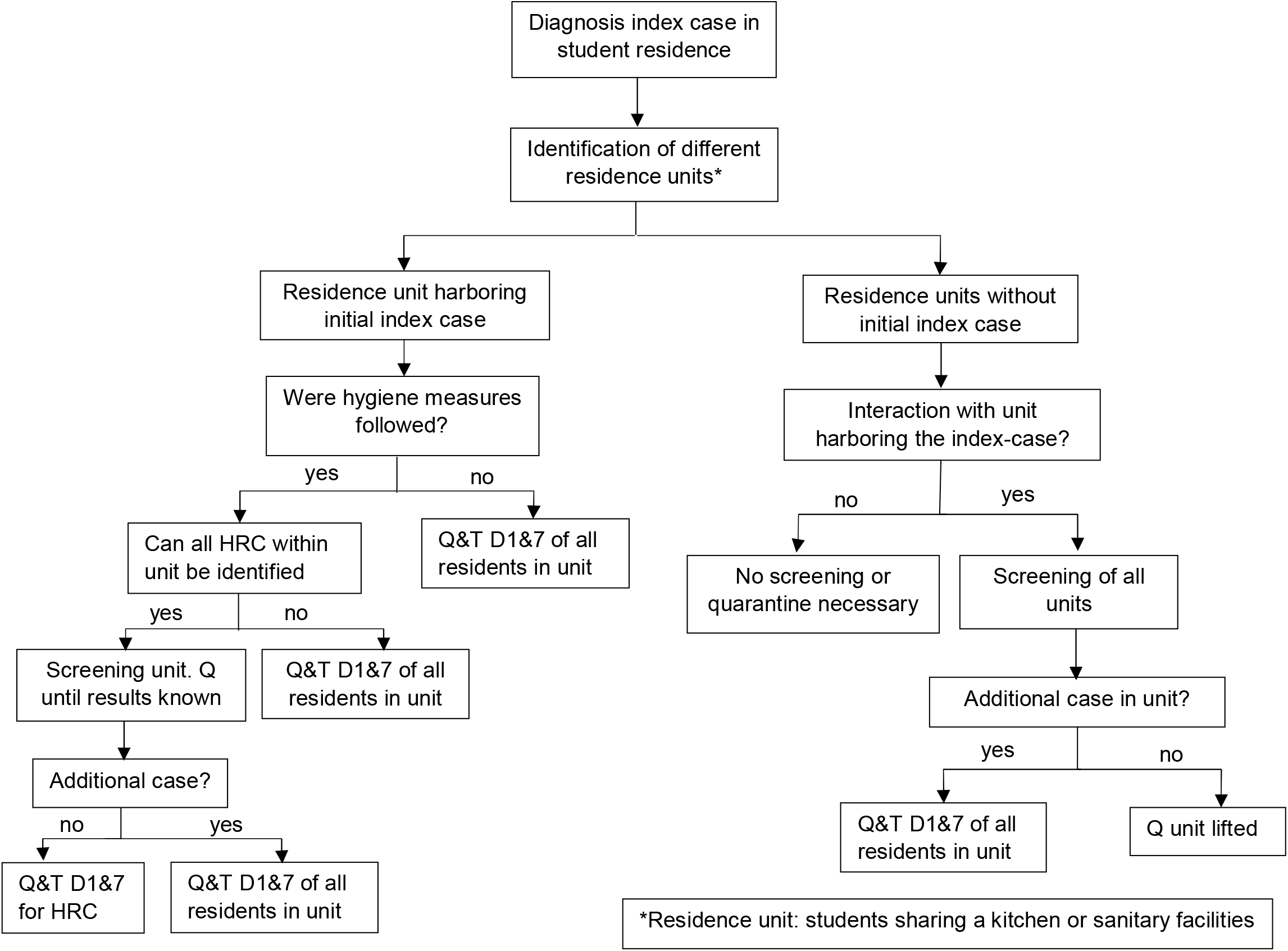
Screening algorithm during a possible student residence outbreak. Abbreviations: D1 = day one, as soon as possible after the diagnosis of the index case. D7 = Day seven, seventh day after the day of diagnosis of the index case, Q = Quarantine, T = testing, HRC = high-risk contact. Once an index case was found to have recently resided in a student residence, all who were part of the residence unit (=students sharing either the same kitchen or the same sanitary facilities) harboring the index case were invited for testing as soon as possible. A target group of students in this residence unit was additionally asked to quarantine and undergo a second test on day 7. The selection of students in this target group depended on whether hygiene measures were strictly complied with and whether high-risk contacts in the strict sense (contact for > 15′ at < 1,5m without face masks, or direct physical contact) could be identified. When regular interaction occurred between the unit harboring the index case and others, those other units were invited for a first round of testing as well. If additional cases were identified in a particular unit, students who belonged to this residence unit were asked to quarantine and undergo a second test on day seven. The detection of new cases in a unit could be responded to by additional screening rounds according to the same protocol

**Figure 2:**
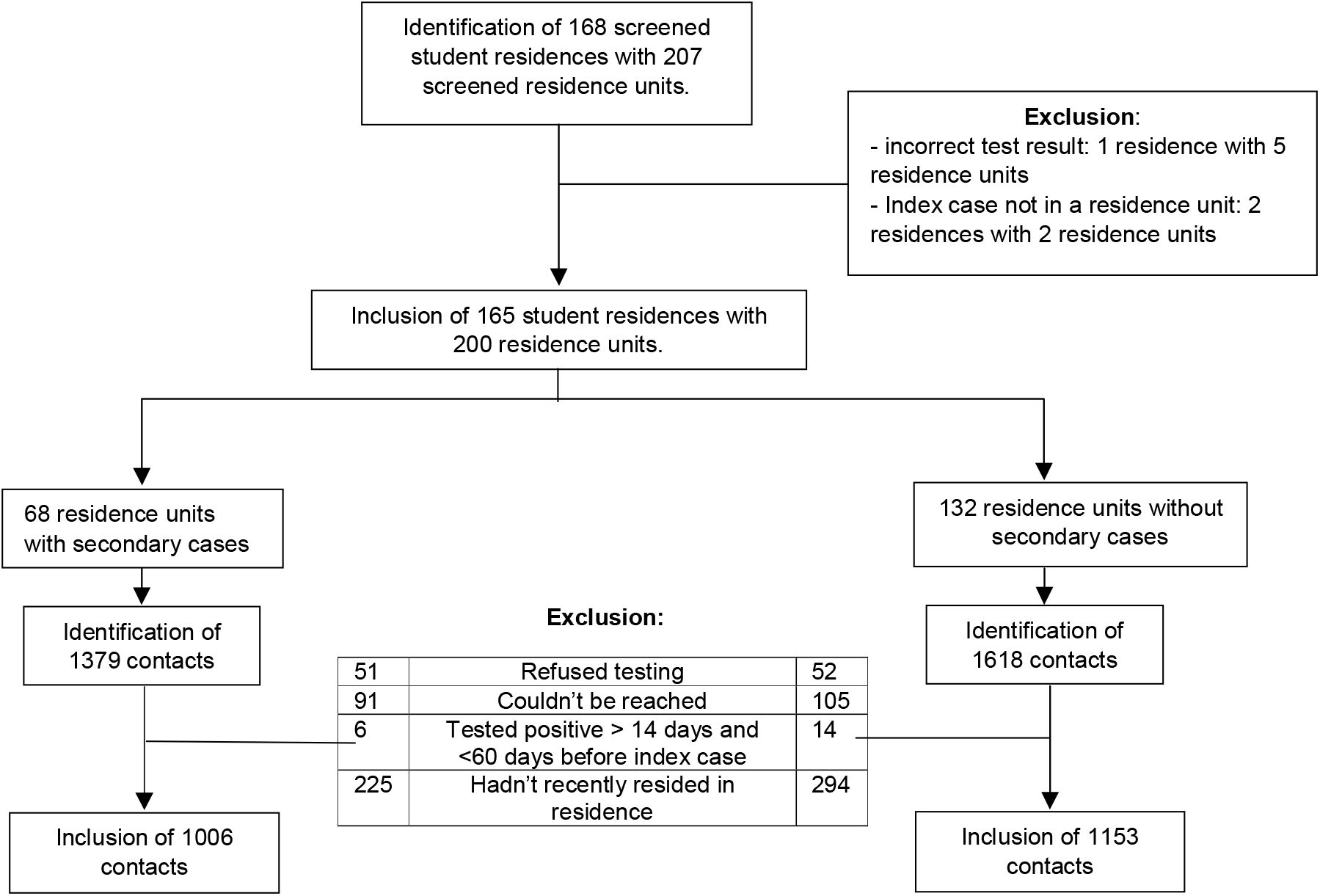
Inclusion and exclusion of student residences and contacts. We included as contacts, all students who resided in a student residence in which a new case of COVID-19 was detected if they met the criteria for further testing as described above. We excluded contacts who were lost to follow up since they could not be reached or refused testing, and contacts who were already diagnosed with COVID-19 up to sixty days but no less than fourteen days before the diagnosis of the index case. We also excluded contacts who reported not having resided in their student residence during the week leading up to the onset of symptoms or diagnosis of the index case, whichever came first.

### Outcome variables

For each student residence, we labelled the case who was diagnosed with COVID-19 by antigen test or a RT-qPCR first as the index case. Contacts included were labelled positive if they were diagnosed with COVID-19 by antigen test or a RT-qPCR in the 2 weeks following and negative if they underwent only a negative test in the 2 weeks following their last contact with the index case or the residence. The SAR in a residence unit was defined as the number of secondary cases in the unit divided by the total number of contacts tested.

### Covariates

We examined the impact of the following covariates on the SAR in a residence unit:

- The delay time between onset of symptoms in the index case and screening of the residence: <4 days delay, ≥ 4 days delay.
- Whether or not the index case shared a kitchen with other residents.
- Whether or not the index case shared sanitary facilities with other residents.
- Whether or not the index reported being infected by a source external to the residence. If the index tested positive during quarantine after traveling abroad, traveling was the external source.
- Whether or not the index attended a social gathering in the residence in the 7 days prior to onset of symptoms or diagnosis characterized by at least two of the following: crowding (at least five individuals belonging at least two households), close contact (<1.5 meters, without the use of face masks) and closed environment (indoor).

In a sensitivity analysis focusing only on the residence units harboring the index case, we distinguished both types of units by adding the binary parameter ‘index case present in the residence unit’.

In 7 out of 200 units screened, two students residing in the same residence tested positive on the same day and were both considered index cases. One of both possible index cases was counted as a secondary case when determining the SAR. An ‘OR’ logic was used for determining the labels of the covariates, meaning it sufficed if one of both index cases reported the presence of the covariate to classify the variable as ‘present’. When the 2 index cases were part of a different residence unit, both of these residence units were classified as units with an index case present.

### Statistical analysis

We analyzed the impact of our covariates on the probability of secondary infections by means of logistic regression while correcting for within-unit and -outbreak correlation. We used generalized estimating equations (GEE) for our primary analysis, which describes the average SAR one may expect when screening a residence in the presence of certain covariate levels. Generalized linear mixed model (GLMM) were used as a validation method. Backward elimination was performed to establish a model in which only significant effects remained. We further performed a sensitivity analysis looking only at the units harboring the initial index case in the residence. Detailed information on the statistical methods used can be found in supplementary materials.

### Study oversight

The Ethics Committee Research UZ/KU Leuven approved our study protocol (study number S64919). The planning, conduct and reporting of the study was performed in line with the Declaration of Helsinki, as revised in 2013. The need for individual informed consent was waived by the Ethics Committee since data was gathered in the context of an ongoing public health response.

## RESULTS

### Included participants

Of the 168 student residences representing 207 residence units located in Leuven or boroughs that were screened between the 30th of October 2020 and the 25th of May 2021 following the detection of at least one confirmed case by antigen or qPCR testing, 165 were included. Three residences representing 7 units were excluded since the index reported not being part of a residence unit. In the remainder, we identified 2997 contacts who met the criteria for screening. We excluded 838 of these contacts due to refusal of testing (103), impossibility to be reached by the contact tracing team (196), recent absence in the student residence (519) and previous infection with COVID-19 (20) leaving 2159 contacts for inclusion in the analysis (Figure 2). The distribution of outbreaks over time can be observed in Supplementary figure 1.

### Student residence characteristics

The main characteristics of the residence units are presented in Table 1.

**Table 1.**
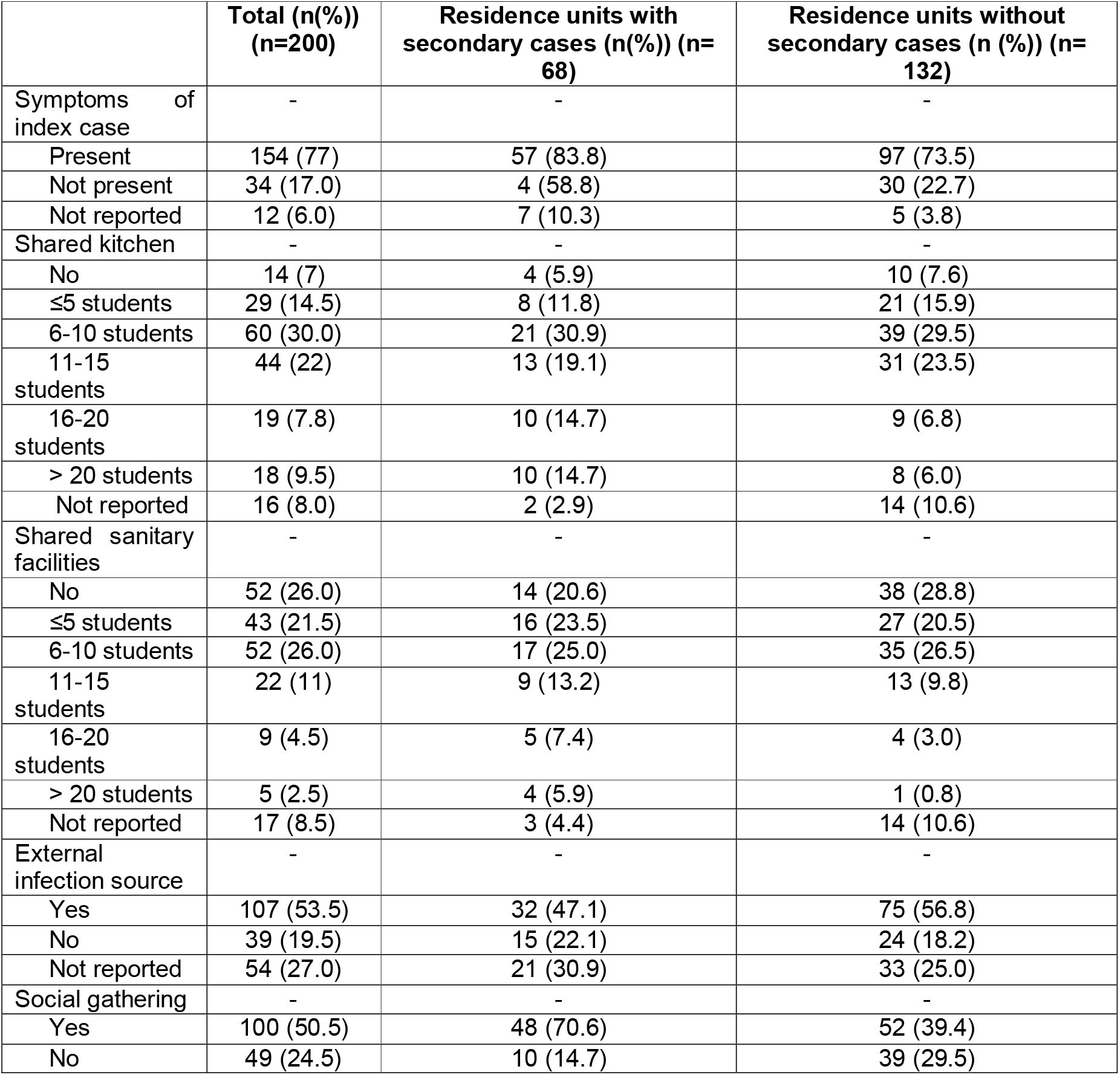
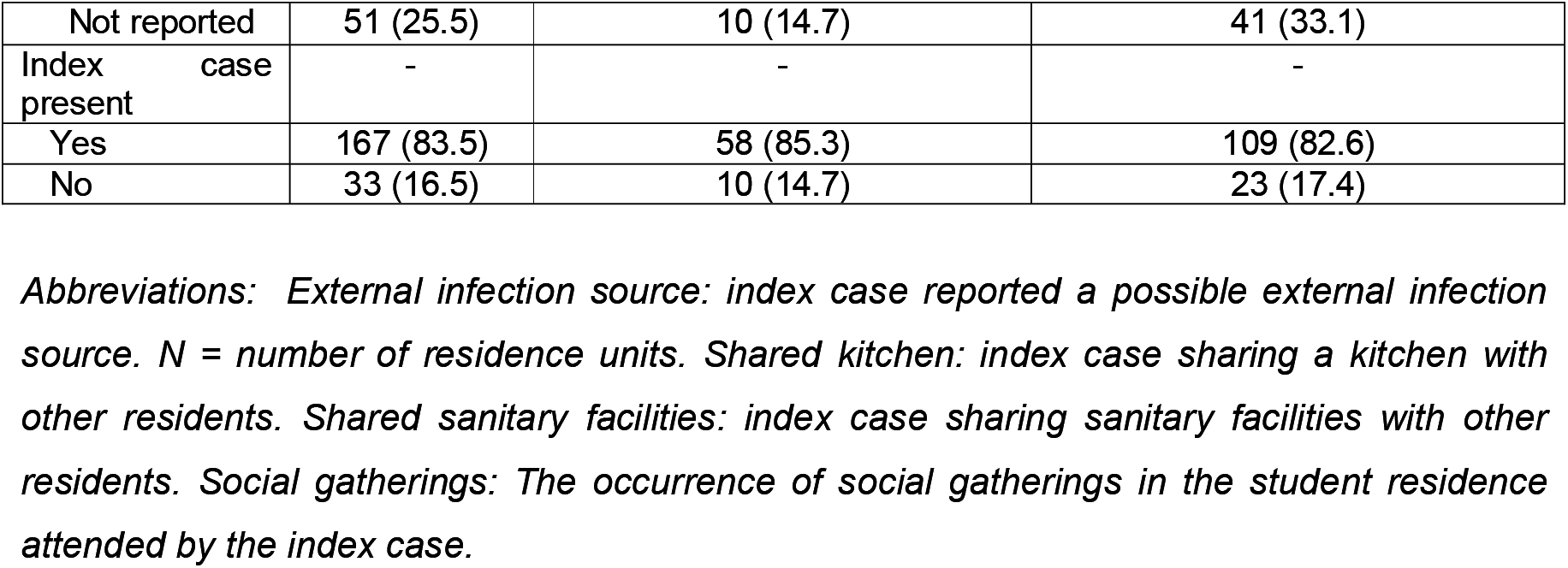
Characteristics of residence units.

|  | **Total (n(%))**<br>**(n=200)** | **Residence units with**<br>**secondary cases (n(%)) (n=68)** | **Residence units without**<br>**secondary cases (n (%)) (n=132)** |
| --- | --- | --- | --- |
| Symptoms of index case | - | - | - |
| Present | 154 (77) | 57 (83.8) | 97 (73.5) |
| Not present | 34 (17.0) | 4 (58.8) | 30 (22.7) |
| Not reported | 12 (6.0) | 7 (10.3) | 5 (3.8) |
| Shared kitchen | - | - | - |
| No | 14 (7) | 4 (5.9) | 10 (7.6) |
| ≤5 students | 29 (14.5) | 8 (11.8) | 21 (15.9) |
| 6-10 students | 60 (30.0) | 21 (30.9) | 39 (29.5) |
| 11-15 students | 44 (22) | 13 (19.1) | 31 (23.5) |
| 16-20 students | 19 (7.8) | 10 (14.7) | 9 (6.8) |
| > 20 students | 18 (9.5) | 10 (14.7) | 8 (6.0) |
| Not reported | 16 (8.0) | 2 (2.9) | 14 (10.6) |
| Shared sanitary facilities | - | - | - |
| No | 52 (26.0) | 14 (20.6) | 38 (28.8) |
| ≤5 students | 43 (21.5) | 16 (23.5) | 27 (20.5) |
| 6-10 students | 52 (26.0) | 17 (25.0) | 35 (26.5) |
| 11-15 students | 22 (11) | 9 (13.2) | 13 (9.8) |
| 16-20 students | 9 (4.5) | 5 (7.4) | 4 (3.0) |
| > 20 students | 5 (2.5) | 4 (5.9) | 1 (0.8) |
| Not reported | 17 (8.5) | 3 (4.4) | 14 (10.6) |
| External infection source | - | - | - |
| Yes | 107 (53.5) | 32 (47.1) | 75 (56.8) |
| No | 39 (19.5) | 15 (22.1) | 24 (18.2) |
| Not reported | 54 (27.0) | 21 (30.9) | 33 (25.0) |
| Social gathering | - | - | - |
| Yes | 100 (50.5) | 48 (70.6) | 52 (39.4) |
| No | 49 (24.5) | 10 (14.7) | 39 (29.5) |
| Not reported | 51 (25.5) | 10 (14.7) | 41 (33.1) |
| Index case present | - | - | - |
| Yes | 167 (83.5) | 58 (85.3) | 109 (82.6) |
| No | 33 (16.5) | 10 (14.7) | 23 (17.4) |

*Abbreviations: External infection source: index case reported a possible external infection source. N = number of residence units. Shared kitchen: index case sharing a kitchen with other residents. Shared sanitary facilities: index case sharing sanitary facilities with other residents. Social gatherings: The occurrence of social gatherings in the student residence attended by the index case.*

Secondary transmission occurred in 68/200 residence units (178 secondary cases), while 132/200 residence units showed no secondary transmission. The median number of secondary cases in the former was 2 (IQR 1-3). The number of negatively screened students ranged between 1 and 103 per residence unit with a median of 7.5 (IQR 5-11). The overall observed secondary attack rate was 8.1% (176/2159).

Symptoms were present in the index case in 77% (154/200) of residence units. The delay time between the onset of symptoms and the follow-up of the residence was shorter than 4 days in 44.5% (89/200) and longer than 4 days in 29.5% (59/200). In most residence units, the index case shared a kitchen with other students (83.8% (170/200)), most frequently with between 6 and 10 other students. The index case also shared sanitary facilities in most residence units (65.5% (131/200)), again most frequently with between 6 and 10 students. In about half of the residence units, the index case reported a possible source of infection outside the student residence. In half of the residence units, the index case reported having attended a social gathering in the student residence in the week before their onset of symptoms or diagnosis. The initial index case of the residence outbreak resided in a residence unit in 83.5% of cases (167/200), which means 16.5% (33/200) of residence units were additionally screened due to interaction between units. No demographics were collected on index cases or contacts. However, the mean and median age of all students tested in the university’s test center during the period under study – who represent the same target population – were 23 and 22, respectively.

### GEE on all residence units

Using a GEE model, the average probability of secondary infections being detected following screening according to the protocol was estimated at 0.0813 (95%CI 0.0705-0.0936), i.e., an estimated SAR between 7.1% and 9.4%. Of the 5 covariates assessed, 3 were removed through backward elimination because they were found to not significantly influence secondary transmission: the delay between symptom onset in the index case and screening of the residence, the shared use of a kitchen and the fact that they had a known source outside of the residence. This left the shared use of sanitary facilities (p=0.04) and the occurrence of a social gathering in the student residence attended by the index case (p=0.0033) as the sole statistically significant predictors for secondary transmission, as presented in table 2 and figure 3. The SAR was lowest at 3% (95%CI 1.5-5.2) for the residence units without the occurrence of a social gathering attended by the index case and without the shared used of sanitary facilities. It was highest at 13% (95%CI 11.4-15.8) in residence units with both risk factors present. An interim position was occupied by units with one risk factor present.

**Table 2:**
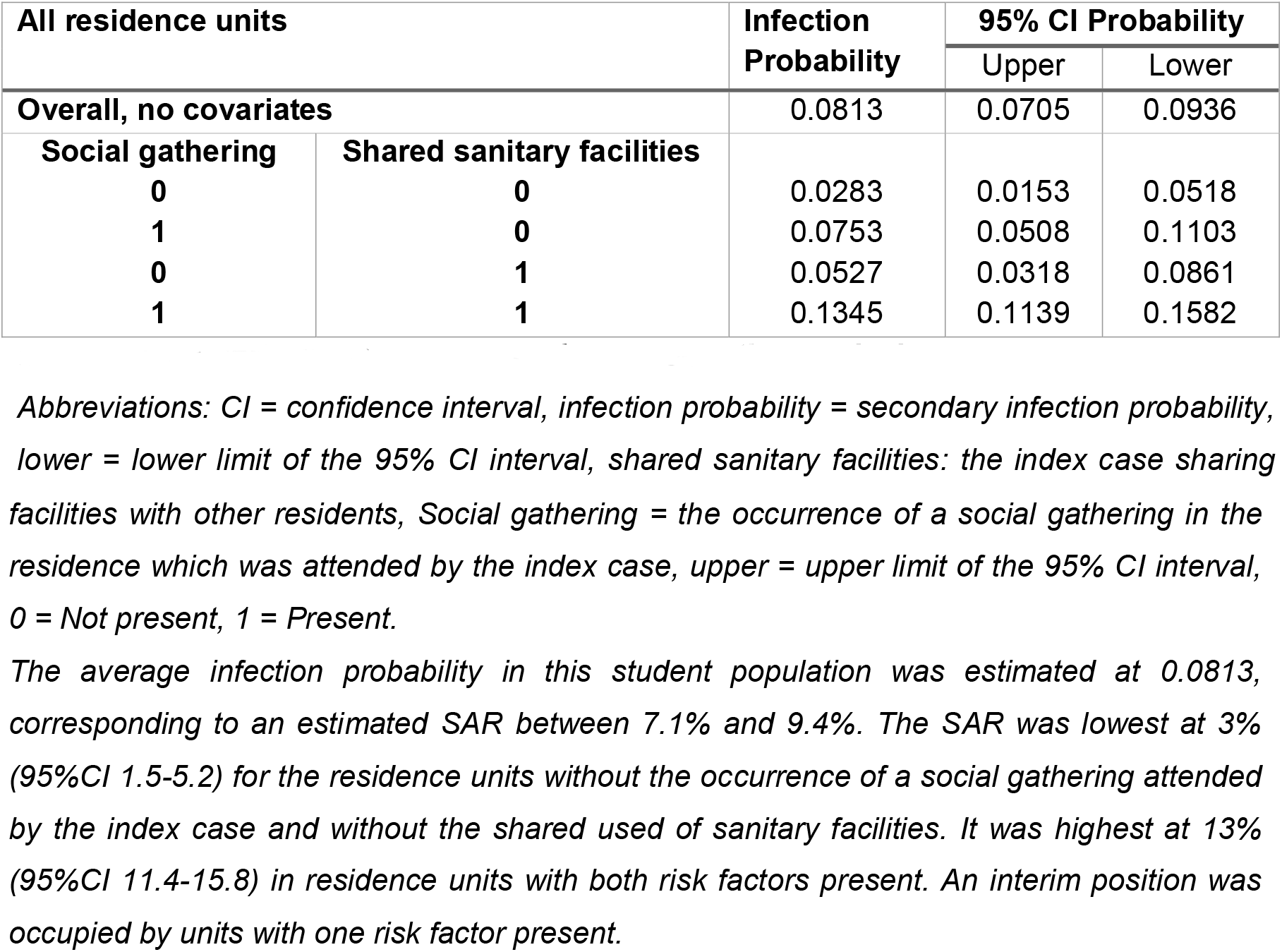
Infection probabilities with confidence intervals and influence of covariates in all residence units by generalized estimated equation analysis.

| All residence units |  | Infection Probability | 95% CI Probability |  |
| --- | --- | --- | --- | --- |
|  |  |  | Upper | Lower |
| Overall, no covariates |  | 0.0813 | 0.0705 | 0.0936 |
| Social gathering | Shared sanitary facilities |  |  |  |
| 0 | 0 | 0.0283 | 0.0153 | 0.0518 |
| 1 | 0 | 0.0753 | 0.0508 | 0.1103 |
| 0 | 1 | 0.0527 | 0.0318 | 0.0861 |
| 1 | 1 | 0.1345 | 0.1139 | 0.1582 |

*Abbreviations: CI = confidence interval, infection probability = secondary infection probability, lower = lower limit of the 95% CI interval, shared sanitary facilities: the index case sharing*
facilities with other residents, Social gathering = the occurrence of a social gathering in the residence which was attended by the index case, upper = upper limit of the 95% CI interval, 0 = Not present, 1 = Present.
The average infection probability in this student population was estimated at 0.0813, corresponding to an estimated SAR between 7.1% and 9.4%. The SAR was lowest at 3% (95%CI 1.5-5.2) for the residence units without the occurrence of a social gathering attended by the index case and without the shared use of sanitary facilities. It was highest at 13% (95%CI 11.4-15.8) in residence units with both risk factors present. An interim position was occupied by units with one risk factor present.

**Figure 3:**
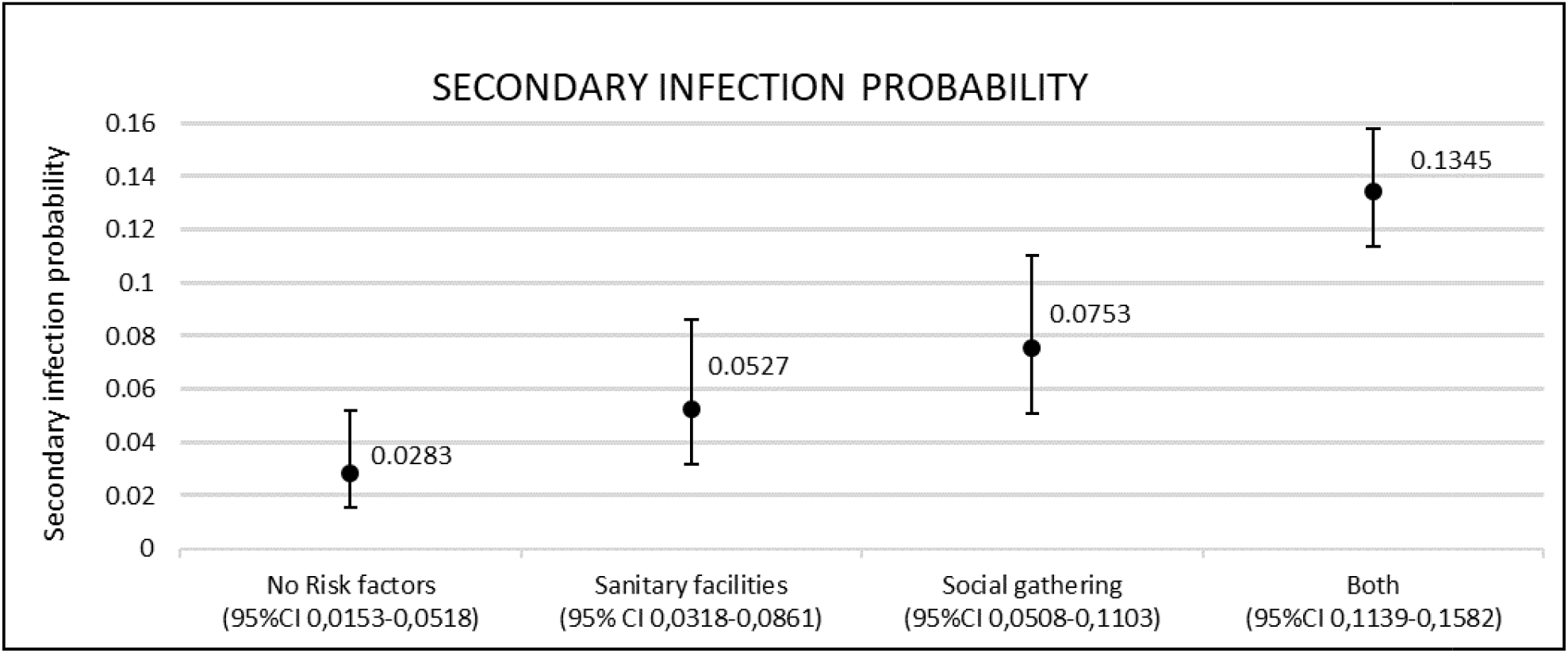
Secondary infection probabilities based on presence of risk factors with 95%CI intervals. Abbreviations: CI = confidence interval. Sanitary facilities = index case sharing sanitary facilities with other residents. Social gathering = occurrence of a social gathering attended by the index case. Both the shared use of sanitary facilities and the occurrence of a social gathering were found to have a significant impact on the secondary infection probability. The SAR was lowest in the group without any risk factors and increased by 10.6% when both were present.

To validate these findings, a GEE model was built using the subset of residence units harboring the initial index case. The same significant covariates were found and they had a similar impact on estimated SAR. The only difference in results we could observe in this analysis was a larger effect of the presence of shared sanitary facilities than when all residence units were analyzed (SI statistical analysis).

### GLLM on all residence units

Using the GLMM model, the average probability of secondary infection was estimated at 0.0855, close to the GEE result, as it should. The 95% confidence interval in this model corresponds to the estimated range of SAR which are to be expected in individual residence units by allowing for a random effect. This was estimated between 0% and 47%. The occurrence of a social gathering remained a significant risk factor (p=0.0019). The shared use of sanitary facilities was borderline non-significant in this model (p=0.0877). However, it was retained for coherence between both approaches. These covariates had a similar impact on the SAR, as presented in table 3 with the range of SAR estimated at 0-17% when both risk factors were present compared to 1-57% when they were absent. A GLMM model on residence units harboring the initial index case revealed similar covariate weights and SAR (SI statistical analysis).

**Table 3:**
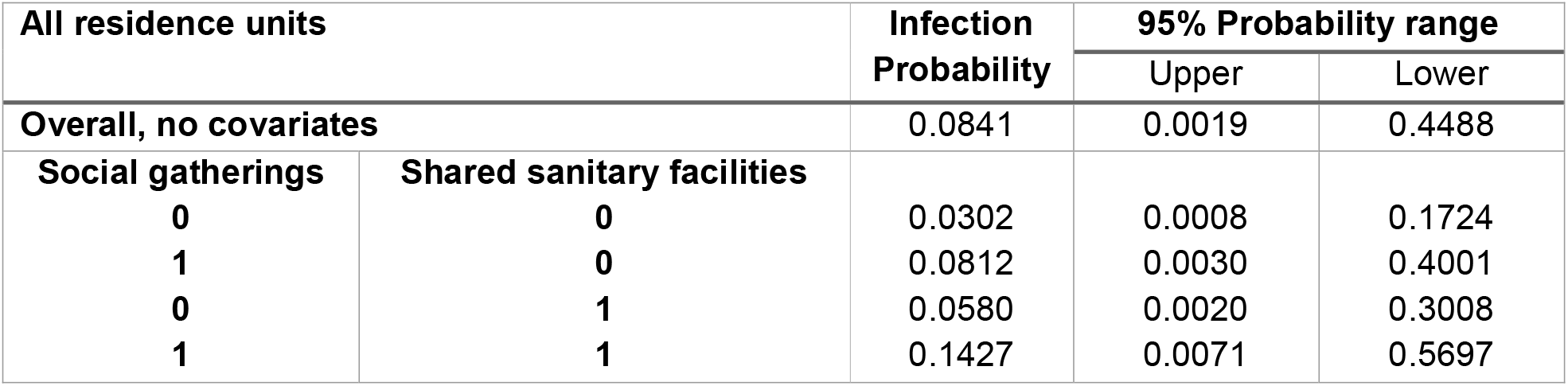
Infection probabilities with 95% probability range and influence of covariates in all residence units by generalized linear mixed model analysis. The 95% probability range corresponds to the estimated range of expected SAR in individual residence units. Abbreviations: Infection probability= secondary infection probability, Lower = Lower limit of the 95% probability range, Shared sanitary facilities: the index case sharing facilities with other residents, social gatherings: the occurrence of a social gathering in the residence, which was attended by the index case. Upper = upper limit of the 95% probability range. 0 = Not present. 1 = Present. The average probability of secondary infection was estimated at 0.0855. with a range of SAR between 0 and 44%. the range of SAR was estimated at 0-17% when both risk factors were present compared to 1-57% when they were absent.

## DISCUSSION

In this large case-ascertainment study, we examined the SAR of COVID-19 infections in a cohort of 165 student residences representing 200 living units and 2326 tested individuals. A standardized risk-based screening protocol was used in all instances following the diagnosis of a first index case in the residence.

Our results show that the overall SAR in student residence units is estimated to lie between 7% and 10%, which is lower than the SAR observed in Belgian households during a similar study period^14^ and other household transmission studies during the alpha dominant pre-vaccination era^15,16^. The results are in line with the 7.8% SAR observed in student residences in a study taking place in a similar setting^17^. The much higher average SAR 27.5% in a Japanese study is based on secondary transmission occurring exclusively in one out of three student dormitories included^7^.

Additionally, we evaluated the influence of 5 risk factors on the overall SAR observed when screening a residence unit, two of which were found to significantly influence the risk of transmission, leading to a SAR that ranged from 3% (95%CI 1.5-5.2) in the absence of either risk factor to 13% (95%CI 11.4-15.8) in the presence of both as assessed by a generalized estimating equations model.

First, the occurrence of an indoor crowded social gathering in the student residence attended by the index case was observed to increase onward transmission, corroborating previous findings pointing at the risk of attending high-risk indoor activities^11,18,19^ and pointing out the importance of indoor social events characterized by crowding and close contact in sparking onward transmission.

Second, the shared use of sanitary facilities significantly increased the probability of identifying secondary cases during screening. This association was found to be a significant factor for acquiring COVID-19 in univariate analysis in one other study. While sanitary facilities were shared in 74% of residence units, preventive measures dissuaded students from concurrent use during the study period. As separations generally exist between installations, this association cannot be explained by droplet transmission. This leaves aerosol-transmission, indirect fomite transmission, fecal-oral and fecal-aerosol transmission as possible underlying mechanisms^20–22^. Alternatively, it could imply that shared use of sanitary facilities constitutes a proxy for overall exposure to an index case living in the same residence unit.

Three factors were not found to be significant, namely whether the index case shared a kitchen, whether they had a known source of infection outside of the residence and whether the delay between onset of symptoms in the index case and the screening of the residence was <4 days or ≥4 days. For any of these factors, a lack of power may be at play. With regards to the sharing of a kitchen, the fact that a larger group of individuals generally shared a kitchen than does a bathroom (Table 1) may imply that the overall exposure to other residents using the same kitchen is generally low. Alternatively, building characteristics or preventive measures in place during the study period may have played a role as well. The fact that the presence of a clear external source of infection in the index case does not significantly influence the SAR within the residence implies that this criterion cannot be used to abstain from screening the residence after diagnosis of a case of COVID-19. While a long delay between symptom onset in the index case and the screening of the unit was not significant, the p-value of 0.0691 in the initial multivariate GEE model for all residence units does convey a trend (see SI).

Sensitivity analyses restricted to residence units harboring the initial index case show that the presence or absence of the initial index case in a residence unit had no significant impact on our conclusions. However, only 17% of the included residence units did not harbor the initial index case and thus the proportion might be too small to draw any firm conclusions. Additionally, residence units which did not harbor the initial index case were only screened and included in this study if there was reported interaction with the residence unit harboring the initial index case. While our analysis corrected for correlation within the same outbreak, further research assessing infection probability in all residence units in a student residence, regardless of interaction between units, can better elucidate the full impact of this parameter.

Our results provide valuable insights into how risk factors assessed in the first index case of COVID-19 in a student residence can inform the subset of residents to be screened thereafter. The crude SAR we observed was high, at 7% to 10%, even though our screening protocol was much broader than screening only contacts with a direct exposure to the first index case. The risk factors found to significantly influence the SAR in the current study provide an evidence base for informed decision making on which subset of individuals should undergo screening in a student residence, thereby improving the balance between comprehensiveness and proportionality of the deployed strategy. Our GLMM analysis demonstrates, however, that a large variation of SAR can still be found even when taking into consideration the risk factors identified, which is the inherent result of the overdispersion associated with COVID-19.

Also, our results improve the evidence base for implementing preventive measures. The importance of shared sanitary facilities is – in the light of the scant and circumstantial evidence base for fecal-oral transmission – rather suggestive of the importance of ventilation for limiting transmission. This fact is also underscored by the increased SAR we observed if a social event had taken place at the residence prior to diagnosis of the first case.

As student residences have many characteristics in common with other collective households, our results have implications for the screening and prevention measures in curative and residential care settings, correctional facilities and the like.

Finally, the fact that social events seem to spur onward transmission implies the need for broader screening of attendants of events characterized by crowding, close contact and closed environment regardless of where this venue may have taken place.

## LIMITATIONS

Our study has several limitations. Firstly, the self-reported nature of most of our data may be subject to recall and reporting bias. Secondly, the population we examined was almost entirely unvaccinated and we did not consider natural immunity in cases or contacts. Thirdly, the alpha variant-of-concern was the dominant variant involved in most of the outbreaks. Fourth, significant general contact restrictions were in place during the study^23^. Fifth, additional parameters likely to influence transmission, such as compliance with preventive measures and detailed building characteristics, were not assessed. Sixth, our analyses were performed on a residence unit level, but not all analyzed parameters were equal for all students in one residence unit. Further research assessing the risk of infection in contacts at the personal level is thus warranted.

## CONCLUSIONS

In this study, we investigated the association of different site- and behavior-specific characteristics which can be elucidated prior to a screening effort on the secondary infection probability of COVID-19 in student residences. Our results show that both the occurrence of an indoor crowded social gathering in the residence unit, which was attended by the index case, and the shared use of sanitary facilities lead to a significant increase in the SAR in the residence unit. While these parameters do not explain all variability in SAR observed in student residence outbreaks, they are nevertheless important factors to be considered when implementing preventive measures, when designing a comprehensive yet proportional testing strategy for student residences and other outbreak-prone environments and when architecturally designing student residences and other shared households.

## Supporting information

Supplementary material including statistical analysis

## Data Availability

All data produced in the present study are available upon reasonable request to the authors

