## Supplementary material including statistical analysis for "Risk factors for SARS-CoV-2 transmission in student residences: a case-ascertained study in Leuven, Belgium from October 2020 until May 2021"

**S1: STATISTICAL ANALYSIS – SECONDARY ATTACK RATE**

### 1 **Data and Methods**

We aim to examine the positivity rate in a unit, with unit defined by ‘unique identifier.’ Positivity is defined as the number of secondary cases divided by the total (secondary cases plus number screened). Of course, the event of an infection is likely correlated for members of the same unit, not only because of shared behaviour, but also due to the residence environment being more or less conducive to spread (e.g., superspreading conditions, such as the formation of aerosols). In addition, in some cases several unique identifiers exist for a common outbreak. It is prudent to allow for correlation between members associated with the same outbreak.

In particular, we aim to explore the impact of a set of covariates on the probability of secondary infections:

- *E_ij_*: external source;
- *H_ij_*: participation in high risk activity;
- *K_ij_*: shared kitchen;
- *S_ij_*: shared sanitary facilities,
- *D*˜*ij* : difference between the date of symptom onset and the date at which follow up started,

with *i* the outbreak unit and *j* the unique identifier within unit *i*.

*D*˜*ij* is expressed as time in days when both dates are observed and missing otherwise. The models will include a categorical variable defined as:

0	if

*D*˜*ij*

is missing (52 cases)

*Dij* =	1	if

2	if We finally define two dummy variables:

| *D*1*,ij* = 1 | if | *D_ij_* = 1*,* |
| --- | --- | --- |
| *D*2*,ij* = 1 | if | *D_ij_* = 2*.* |

*D*˜*ij <* 4 (70 cases)

*D*˜*ij* ≥ 4 (79 cases)

Denote by *Y_ijk_* = 1 that an infection is observed and *Y_ijk_* = 0 otherwise. Here, *k* ranges over the individuals within unit *j*.

1.1 **Modeling Frameworks and Models**

It is natural to model the probability of secondary infection, accounting for covariates, by means of logistic regression, but with a correction for within-unit and -outbreak correlation. This will be done in two distinct and complementary ways.

The first approach is **generalized estimating equations**, where a so-called working correlation is added to the model. While the logistic regression parameters retain their usual interpretation, this approach ensures that valid parameter and precision estimates are obtained, even though within-unit data are dependent.

The logistic expression takes the form, for example when all covariates are present:

*P* (*Y_ijk_* = 1|*E_ij_, H_ij_, K_ij_, S_ij_*)

exp(*β*_0_ + *β*_1_*E_ij_* + *β*_2_*H_ij_* + *β*_3_*K_ij_* + *β*_4_*S_ij_* + *β*5*D*_1_*_,ij_* + *β*_6_*D*_2_*_,ij_*)

=	*.*	(1)

1 + exp(*β*_0_ + *β*_1_*E_ij_* + *β*_2_*H_ij_* + *β*_3_*K_ij_* + *β*_4_*S_ij_* + *β*5*D*_1_*_,ij_* + *β*_6_*D*_2_*_,ij_*)

While (1) represents a specific instance, (2) is a generic expression, where all covariates are grouped into a vector *x_ij_* and all parameters into a vector *β*:

exp(*x’*^'^*β*)

*ij*

*P* (*Y_ijk_* = 1|*x_ij_*) =

*ij*

1 + exp(*x’*^'^*β*)

*.*	(2)

Fitting the model produces parameter estimates, standard errors, and *p*-values. The latter will be used to perform backward selection, where in a stepwise fashion the least significant effects are removed until only significant effects remain. Apart from a model with covariates, one with only an intercept will be fitted, to estimate the overall probability of infection.

Given parameter estimates, (2) can be used to estimate the corresponding probabilities. In order to obtain standard errors and confidence intervals for these probabilities, the so-called delta method is used, as follows. Denote, for simplicity, for a given unit (which can but does not have to belong to the actual sample) *p* = *P* (*Y_i_*_0_ *j*_0_ *k* = 1|*x, β*), then

'

var(*p*) $\cong$*x’ p*(1 − *p*)var(*β*)*p*(1 − *p*)*x,*

where (*i*_0_*, j*_0_) refers to either an existing or an artificially composed unit, and where var(*β*) is the

variance-covariance matrix of the parameter estimates, routinely available from statistical software (e.g., SAS or R). Confidence intervals are obtained by first constructing a 95% confidence interval for the linear predictor *η* = *x’*^'^ *β*, say [*η_L_, η_U_* ], and then calculating the corresponding [*p_L_, p_U_* ] via expit formula (2).

*ij*

The second approach is a **generalized linear mixed model**), where a random effect is included to account for the within-unit association:

*P* (*Y_ijk_* = 1|*E_ij_, H_ij_, K_ij_, S_ij_, b_i_*)

exp(*γ*_0_ + *b_i_γ*_1_*E_ij_* + *γ*_2_*H_ij_* + *γ*_3_*K_ij_* + *γ*_4_*S_ij_*) + *γ*_5_*D*_1_*_,ij_* + *γ*_6_*D*_2_*_,ij_*

=	*,*	(3)

1 + exp(*γ*_0_ + *b_i_* + *γ*_1_*E_ij_* + *γ*_2_*H_ij_* + *γ*_3_*K_ij_* + *γ*_4_*S_ij_* + *γ*_5_*D*_1_*_,ij_* + *γ*_6_*D*_2_*_,ij_*)

with *b_i_* ∼ *N* (0*, d*). Also here, a generic version can be written down:

exp(*x’*^'^ *γ* + *b_i_*)

*ij*

*P* (*Y_ijk_* = 1|*x_ij_*)	=

*ij*

1 + exp(*x’*^'^ *γ* + *b_i_*)

*,*	(4)

The *γ* parameters should not be seen as alternative estimates for their *β* counterparts. Whereas (1) can be used to estimate the average probability of infection, given levels for the various covariates, model (3) describes the probability of infection, given covariate levels but also given a certain level of random effect *b_i_*. The use of the model is that it easily allows to describe the typical range of infection rates that one sees across units with the same covariate levels.

A simple, sampling-based method to obtain this range is by sampling a large number of copies from a *N* (0*, d*) distribution, calculating for each the corresponding probability, and then deriving the desired quantities, such as their 2.5% and 97.5% percentiles. Evidently, also the mean or median can be derived.

All analyses will be done in two versions. In the first version, only units with index case present are analysed. In the second version, all units will be analysed together, with or without index case present (and allowing the presence or absence of an index case as an additional predictor).

1.2 **Missing Data**

The GLMM is likelihood-based, implying that it is valid under the assumption of missing at random, which means that missingness may depend on outcomes (both *X* and *Y* variables) in as far as they are observed but, given these, not further on unobserved outcomes. GEE has a frequentist basis and therefore is generally valid only under the stronger assumption of missing completely at random, meaning that missingness can depend on observed covariates (*X* variables) but not on *Y* variables. Given that we will compare the GLMM and GEE results, we can compare the results of both, in terms of model-based attack rates; if these are similar, the same level of confidence can be placed in our GEE results as in our GLMM results.

### 2 **Analysis of All Clusters, With or Without Index Case Present**

2.1 **Models Without Covariates**

Using GEE, the following estimate is obtained:

GEE, no covariates, all clusters

Effect	Par.	Estimate	(s.e.)	*p*

Intercept	*β*_0_	-2.4244	(0.1481)	—

The intraclass correlation is *α* = 0*.*038. This seems surprisingly low, but arguably the correlation structure is much more complex than a simple constant. For GEE, this is not a problem as the intraclass correlation is not of direct interest. The estimates and standard errors are valid, even when the correlation structure is misspecified.

The corresponding average probability of infection is *p* = 0*.*0813 with s.e. 0*.*0059. The confidence interval is [0*.*0705; 0*.*0936].

Turning to the GLMM version, we find:

GLMM, no covariates, all clusters

| Effect | Par. | Estimate | (s.e.) | *p* |
| --- | --- | --- | --- | --- |
| Intercept | *γ*_0_ | -3.2212 | (0.2217) |  |
| Variance R.I. | *d* | 2.3202 | (0.5865) | 0.0001 |

Sampling a 100,000 copies from a *N* (0*, d* = 2*.*3202) distribution, we find that the average probability is 0*.*0841, close to the GEE result, as it should be, with a range [0*.*0019; 0*.*4488].

It is important to emphasize the difference in interpretation between the GEE and GLMM ranges. Taken together, we conclude that the overall secondary attack rate lies somewhere between 7.1% and 9.4% with 95% confidence, and that in individual clusters, we observe attack rates that go from nearly 0% to about 45%.

2.2 **Models With Covariates**
    2.2.1 **GEE Results**

We first present the individual (univariate) effect of all covariates.

GEE, individual covariate effects, all clusters

| Effect | Par. | Estimate | (s.e.) *p* | |
| --- | --- | --- | --- | --- |
| Intercept | *β*_0_ | -2.2508 | (0.2503) | — |
| External source | *β*_1_ | -0.3658 | (0.3394) | 0.2811 |
| Intercept | *β*_0_ | -3.0955 | (0.3191) | — |
| High risk | *β*_2_ | 1.0190 | (0.3625) | 0.0049 |
| Intercept | *β*_0_ | -3.0366 | (0.5188) | — |
| Kitchen | *β*_3_ | 0.7231 | (0.5359) | 0.1772 |
| Intercept | *β*_0_ | -3.2114 | (0.2535) | — |
| Sanitary fac. | *β*_4_ | 1.0327 | (0.2995) | 0.0006 |
| Intercept | *β*_0_ | -2.8415 | (0.3112) | — |
| Day diff. 1 | *β*_5_ | 0.4709 | (0.4033) | 0.2429 |
| Day diff. 2 | *β*_6_ | 0.5353 | (0.3767) | 0.1553 |
| Joint effect Day diff. |  |  |  | 0.3022 |

A model including all covariates at once, which will serve as a basis for backward selection, shows the following results:

GEE, initial multivariate model, all clusters

| Effect | Par. | Estimate | (s.e.) *p* | |
| --- | --- | --- | --- | --- |
| Intercept | *β*_0_ | -5.4686 | (1.1958) | — |
| External source | *β*_1_ | -0.1119 | (0.4089) | 0.7844 |
| High risk | *β*_2_ | 0.6062 | (0.3817) | 0.1122 |
| Kitchen | *β*_3_ | 1.4169 | (0.7631) | 0.0633 |
| Sanitary fac. | *β*_4_ | 0.5867 | (0.3383) | 0.0829 |
| Day diff. 1 | *β*_5_ | 0.7660 | (0.5404) | 0.1563 |
| Day diff. 2 | *β*_6_ | 1.0642 | (0.5854) | 0.0691 |
| Joint effect Day diff. |  |  |  | 0.2791 |

Of all covariates placed into the model, external source, the difference in days, and the presence of a shared kitchen have no significant impact and are removed, in this order. This leaves high risk and sanitary facilities as the sole predictors. An interaction between them is not necessary. The sanitary facilities variable is significant in GEE but will be borderline non-significant in GLMM (see further). We decided to keep it in for coherence between all approaches.

The final GEE parameter estimates after model simplification are:

GEE, covariates, all clusters

Effect	Par.	Estimate	(s.e.)	*p*

| Intercept | *β*_0_ | -3.5362 | (0.4234) | — |
| --- | --- | --- | --- | --- |
| High risk | *β*_2_ | 1.0282 | (0.3505) | 0.0033 |
| Sanitary fac. | *β*_4_ | 0.6464 | (0.3147) | 0.0400 |

The intraclass correlation is now *α* = −0*.*0402.

The variance=covariance matrix of the parameter estimates takes values:

var(*β*) = 

0*.*10277	−0*.*06755	−0*.*04179

−0*.*06755	0*.*07830 −0*.*003146

−0*.*04179 −0*.*003146	0*.*05331

The probability of infection and corresponding confidence interval is now different between covariate levels:

GEE, covariates, all clusters; infection probabilities

| High risk | Sanitary fac. | Infection prob. | (s.e.) | confidence interval |
| --- | --- | --- | --- | --- |
| 0 | 0 | 0.0283 | (0.0088) | [0.0153;0.0518] |
| 1 | 0 | 0.0753 | (0.0149) | [0.0508;0.1103] |
| 0 | 1 | 0.0527 | (0.0134) | [0.0318;0.0861] |
| 1 | 1 | 0.1345 | (0.0133) | [0.1139;0.1582] |

2.1.1 **GLMM Results**

The results for the individual predictors are:

GLMM, individual covariate effects, all clusters

| Effect | Par. | Estimate | (s.e.) *p* | |
| --- | --- | --- | --- | --- |
| Intercept | *γ*_0_ | -3.1539 | (0.3734) | — |
| External source | *γ*_1_ | -0.3556 | (0.4060) | 0.3828 |
| Variance R.I. | *d* | 2.3449 | (0.7085) | 0.0012 |
| Intercept | *γ*_0_ | -3.8647 | (0.4246) | — |
| High risk | *γ*_2_ | 1.3014 | (0.4445) | 0.0041 |
| Variance R.I. | *d* | 2.0066 | (0.5852) | 0.0008 |
| Intercept | *γ*_0_ | -3.5811 | (0.6729) | — |
| Kitchen | *γ*_3_ | 0.4208 | (0.6823) | 0.5282 |
| Variance R.I. | *d* | 2.2421 | (0.5860) | 0.0002 |
| Intercept | *γ*_0_ | -4.0201 | (0.4192) | — |
| Sanitary fac. | *γ*_4_ | 1.1200 | (0.4307) | 0.0102 |
| Variance R.I. | *d* | 2.0108 | (0.5442) | 0.0003 |
| Intercept | *γ*_0_ | -3.7061 | (0.3970) | — |
| Day diff. 1 | *γ*_5_ | 0.5382 | (0.4554) | 0.2389 |
| Day diff. 2 | *γ*_6_ | 0.7664 | (0.4518) | 0.0917 |
| Joint effect Day diff. |  |  |  | 0.1607 |
| Variance R.I. | *d* | 2.2692 | (0.5762) | 0.0001 |

The model with all effects included, prior to model selection, is as follows:

GLMM, initial multivariate model, all clusters

| Effect | Par. | Estimate | (s.e.) *p* | |
| --- | --- | --- | --- | --- |
| Intercept | *γ*_0_ | -6.1100 | (1.2537) | — |
| External source | *γ*_1_ | 0.0524 | (0.4443) | 0.9064 |
| High risk | *γ*_2_ | 0.8160 | (0.4745) | 0.0889 |
| Kitchen | *γ*_3_ | 1.2238 | (0.8792) | 0.1673 |
| Sanitary fac. | *γ*_4_ | 0.7998 | (0.4895) | 0.1057 |
| Day diff. 1 | *γ*_5_ | 0.8348 | (0.6111) | 0.1753 |
| Day diff. 2 | *γ*_6_ | 1.2088 | (0.6086) | 0.0501 |
| Joint effect Day diff. |  |  |  | 0.1447 |
| Variance R.I. | *d* | 1.4790 | (0.5346) | 0.0069 |

Also here, the variables external source, shared kitchen facility, and difference in days are remove, to lead to the following final model:

GLMM, covariates, all clusters

Effect	Par.	Estimate	(s.e.)	*p*

| Intercept | *γ*_0_ | -4.3351 | (0.5365) | — |
| --- | --- | --- | --- | --- |
| High risk | *γ*_2_ | 1.2139 | (0.4267) | 0.0052 |
| Sanitary fac. | *γ*_4_ | 0.8025 | (0.4660) | 0.0877 |
| Variance R.I. | *d* | 1.7072 | (0.5353) | 0.0018 |

The variance=covariance matrix of the parameter estimates takes values:

var(*γ*) =

0*.*2883	−0*.*1345	−0*.*1629

−0*.*1345	0*.*1821 −0*.*01694

−0*.*1629 −0*.*01694	0*.*2172

The mean probability of infection is given, together with the 95% range of infection probabilities among clusters at a given covariate level:

GLMM, covariates, all clusters; infection probabilities and ranges

| High risk | Sanitary fac. | Infection prob. | range |
| --- | --- | --- | --- |
| 0 | 0 | 0.0302 | [0.0008;0.1724] |
| 1 | 0 | 0.0812 | [0.0030;0.4001] |
| 0 | 1 | 0.0580 | [0.0020;0.3008] |
| 1 | 1 | 0.1427 | [0.0071;0.5697] |

Unsurprisingly, the secondary attack rate is smallest for those clusters without high risk activity and without shared sanitary facilities, it is estimates at about 3%, allowing for uncertainty, it is fair to say that it ranges between 2% and 5%. The bulk of the clusters in this category exhibit secondary attack rates between 0% and 17%.

The highest values are obtained for the mirror group, made up of those clusters with both high risk activity and shared sanitary facilities, with attack rate 11%–16%, and the vast majority of individual clusters ranging from 1% to 57%.

The other two groups take an interim position between these two extremes.

### 3 **Analysis of Clusters With Index Case Present Only**

3.1 **Models Without Covariates**

Using GEE, the following estimate is obtained:

GEE, no covariates, clusters with index present

Effect	Par.	Estimate	(s.e.)	*p*

Intercept	*β*_0_	-2.3948	(0.1542)	—

The intraclass correlation is *α* = −0*.*39.

The corresponding probability of infection is *p* = 0*.*0836 with s.e. 0*.*0065. The confidence interval is [0*.*0717; 0*.*0973].

Turning to the GLMM version, we find:

GLMM, no covariates, clusters with index present

| Effect | Par. | Estimate | (s.e.) | *p* |
| --- | --- | --- | --- | --- |
| Intercept | *γ*_0_ | -3.2965 | (0.2422) |  |
| Variance R.I. | *d* | 2.6262 | (0.6731) | 0.0001 |

Sampling a 100,000 copies from a *N* (0*, d* = 2*.*6262) distribution, we find that the average probability is 0*.*0855 with a range [0*.*0015; 0*.*4774].

It is clear that the inclusion or exclusion of those clusters without an index case has virtually no impact on the conclusions.

3.2 **Models With Covariates**

When restricting to those with index case present, results are largely similar, both for GEE as well as GLMM. We present the results in what follows.

3.2.1 **GEE Results**

The individual effects of the covariates, examined in univariate models, are as follows:

GEE, individual covariate effects, clusters with index

| Effect | Par. | Estimate | (s.e.) *p* | |
| --- | --- | --- | --- | --- |
| Intercept | *β*_0_ | -2.2867 | (0.2692) | — |
| External source | *β*_1_ | -0.4813 | (0.3671) | 0.1898 |
| Intercept | *β*_0_ | -3.0880 | (0.3164) | — |
| High risk | *β*_2_ | 1.0640 | (0.3633) | 0.0034 |
| Intercept | *β*_0_ | -3.0366 | (0.5188) | — |
| Kitchen | *β*_3_ | 0.6936 | (0.5429) | 0.2014 |
| Intercept | *β*_0_ | -3.1850 | (0.2883) | — |
| Sanitary fac. | *β*_4_ | 0.9895 | (0.3346) | 0.0031 |
| Intercept | *β*_0_ | -2.7265 | (0.3239) | — |
| Day diff. 1 | *β*_5_ | 0.3100 | (0.4260) | 0.4668 |
| Day diff. 2 | *β*_6_ | 0.5016 | (0.3886) | 0.1967 |
| Joint effect Day diff. |  |  |  | 0.4190 |

A model including all covariates at once, which will also here serve as a basis for backward selection, shows the following results:

GEE, initial multivariate model, clusters with index

| Effect | Par. | Estimate | (s.e.) *p* | |
| --- | --- | --- | --- | --- |
| Intercept | *β*_0_ | -5.7920 | (1.2640) | — |
| External source | *β*_1_ | 0.0825 | (0.5425) | 0.8792 |
| High risk | *β*_2_ | 0.5640 | (0.3831) | 0.1410 |
| Kitchen | *β*_3_ | 1.4388 | (0.8029) | 0.0732 |
| Sanitary fac. | *β*_4_ | 0.9612 | (0.4419) | 0.0296 |
| Day diff. 1 | *β*_5_ | 0.8209 | (0.5458) | 0.1326 |
| Day diff. 2 | *β*_6_ | 1.0212 | (0.7120) | 0.1515 |
| Joint effect Day diff. |  |  |  | 0.3181 |

Like in the case with all clusters involved, the backward selection leads to the removal, respectively of external source, difference in days, and shared kitchen facilities. The final GEE model takes the form:

GEE, covariates, clusters with index

Effect	Par.	Estimate	(s.e.)	*p*

| Intercept | *β*_0_ | -3.8484 | (0.4884) | — |
| --- | --- | --- | --- | --- |
| High risk | *β*_2_ | 0.9796 | (0.3585) | 0.0063 |
| Sanitary fac. | *β*_4_ | 1.0338 | (0.3432) | 0.0026 |

The intraclass correlation is now *α* = 0*.*62. This is noteworthy, as it is the first time that a substantial correlation is detected. Arguably, this is due to the presence of the index with in addition correction for important covariates, so that the residual correlation is easier to estimate.

Note also that the effect of sanitary facilities is more important in this case than when all clusters are analyzed.

The variance-covariance matrix of the parameter estimates takes values:

var(*β*) =

0*.*12837	−0*.*06650	−0*.*07037

−0*.*06550	0*.*08040	−0*.*005700

−0*.*07037 −0*.*005700	0*.*08513

The probability of infection and corresponding confidence interval, for the four levels:

GEE, covariates, clusters with index; infection probabilities

| High risk | Sanitary fac. | Infection prob. | (s.e.) | confidence interval |
| --- | --- | --- | --- | --- |
| 0 | 0 | 0.0209 | (0.0073) | [0.0105;0.0412] |
| 1 | 0 | 0.0537 | (0.0142) | [0.0318;0.0893] |
| 0 | 1 | 0.0565 | (0.0144) | [0.0341;0.0923] |
| 1 | 1 | 0.1376 | (0.0123) | [0.1152;0.1636] |

3.1.1 **GLMM Results**

The results for the individual predictors in this case are:

GLMM, individual covariate effects, only clusters with index

| Effect | Par. | Estimate | (s.e.) *p* | |
| --- | --- | --- | --- | --- |
| Intercept | *γ*_0_ | -3.2841 | (0.4187) | — |
| External source | *γ*_1_ | -0.3635 | (0.4659) | 0.4367 |
| Variance R.I. | *d* | 2.6696 | (0.8091) | 0.0001 |
| Intercept | *γ*_0_ | -3.8985 | (0.4432) | — |
| High risk | *γ*_2_ | 1.2765 | (0.4670) | 0.0072 |
| Variance R.I. | *d* | 2.2049 | (0.6620) | 0.0011 |
| Intercept | *γ*_0_ | -3.6565 | (0.7032) | — |
| Kitchen | *γ*_3_ | 0.4172 | (0.7121) | 0.5588 |
| Variance R.I. | *d* | 2.5524 | (0.6652) | 0.0002 |
| Intercept | *γ*_0_ | -4.0813 | (0.4587) | — |
| Sanitary fac. | *γ*_4_ | 1.0860 | (0.4727) | 0.0229 |
| Variance R.I. | *d* | 2.3208 | (0.6237) | 0.0003 |
| Intercept | *γ*_0_ | -3.6941 | (0.4161) | — |
| Day diff. 1 | *γ*_5_ | 0.4622 | (0.4851) | 0.3420 |
| Day diff. 2 | *γ*_6_ | 0.6303 | (0.1960) | -0.3284 |
| Joint effect Day diff. |  |  |  | 0.2567 |
| Variance R.I. | *d* | 2.5773 | (0.6631) | 0.0001 |

The model with all effects included, prior to model selection, is as follows:

GLMM, initial multivariate model, only clusters with index

| Effect | Par. | Estimate | (s.e.) *p* | |
| --- | --- | --- | --- | --- |
| Intercept | *γ*_0_ | -6.2146 | (1.3004) | — |
| External source | *γ*_1_ | 0.0746 | (0.4913) | 0.8796 |
| High risk | *γ*_2_ | 0.7019 | (0.5022) | 0.1656 |
| Kitchen | *γ*_3_ | 1.2251 | (0.9036) | 0.1786 |
| Sanitary fac. | *γ*_4_ | 0.9344 | (0.5318) | 0.0823 |
| Day diff. 1 | *γ*_5_ | 0.8321 | (0.6443) | 0.1998 |
| Day diff. 2 | *γ*_6_ | 1.1536 | (0.6518) | 0.0802 |
| Joint effect Day diff. |  |  |  | 0.1766 |
| Variance R.I. | *d* | 1.6297 | (0.5957) | 0.0075 |

Once again, the variables external source, shared kitchen facility, and difference in days are remove, to lead to the following final model:

The GLMM results for the final, reduced model are:

GLMM, covariates, clusters with index

Effect	Par.	Estimate	(s.e.)	*p*

| Intercept | *γ*_0_ | -4.5024 | (0.5716) | — |
| --- | --- | --- | --- | --- |
| High risk | *γ*_2_ | 1.1671 | (0.4492) | 0.0106 |
| Sanitary fac. | *γ*_4_ | 0.9752 | (0.5035) | 0.0552 |
| Variance R.I. | *d* | 1.8957 | (0.5967) | 0.0019 |

The variance=covariance matrix of the parameter estimates takes values:

var(*γ*) = 

0*.*3267	−0*.*1435	−0*.*1933

−0*.*1435	0*.*2018 −0*.*01987

−0*.*1933 −0*.*01987	0*.*2535

The mean probability of infection is given, together with the 95% range of infection probabilities among clusters at a given covariate level:

GLMM, covariates, clusters with index; infection probabilities

| and ranges |  | | |
| --- | --- | --- | --- |
| High risk | Sanitary fac. | Infection prob. | range |
| 0 | 0 | 0.0282 | [0.0006;0.1685] |
| 1 | 0 | 0.0733 | [0.0020;0.3897] |
| 0 | 1 | 0.0616 | [0.0017;0.3316] |
| 1 | 1 | 0.1440 | [0.0062;0.5917] |

Comparing these results to the ones where all clusters are analyzed shows the relatively stronger importance of shared sanitary facilities in this case.

### 1 **Concluding Remarks**

Whether or not the clusters without index case present are included changes the conclusions only mildly, although the importance of shared sanitary facilities increases when restricted to units with the index case presnt.

Broadly, the overall secondary attack rate is estimated to lie between 7% and 10%, but individual clusters show considerable variability with the bulk of them exhibiting a rate between 0% and nearly 50%.

When correcting for covariates, high risk activity and shared sanitary facilities are very important.

External source and shared kitchen play no significant role next to these predictors.

When both high risk activity and shared sanitary facilities are present, then the attack rate varies from 1% to nearly 60% for such clusters, whereas it typically remains lower than 17% when both risk factors are absent. When one of the two is present, the attack rate is on average about 3–9% and roughly ranges over 0–35% for individual clusters.

**Supplementary 2: Sensitivity analysis**

A sensitivity analysis was performed analysing only those residence units harbouring an index case. Using the GEE model, results were largely comparable to the results of the analysis of all residence units. The average probability of secondary infections being detected following screening according to the protocol was estimated at 0.0836 (95%CI 0.0717-0.0973), corresponding to an estimated secondary attack rate between 7.2% and 9.7%. Of the 5 covariates assessed, 3 were removed through backward elimination because they were found to not significantly influence secondary transmission: the delay between symptom onset in the index case and screening of the residence, the shared use of a kitchen and the fact that they had a known source outside of the residence. This left the shared use of sanitary facilities (p=0.0026) and the occurrence of a social gathering in the student residence attended by the index case (p=0.0063) as the only statistically significant predictors for secondary transmission, as presented in table 2 and figure 3. The secondary attack rate was lowest at 2²% (95%CI 1.0-4.1) for the residence units without the occurrence of a social gathering attended by the index case and without the shared used of sanitary facilities. It was highest at 14% (95%CI 11.5-16.4) in residence units with both risk factors present. An interim position was occupied by units with one risk factor present. The presence or absence of the index case in the residence unit thus had little influence on our results

| Residence units with index case present | | Infection  Probability | 95% CI Probability | |
| --- | --- | --- | --- | --- |
|  |  |  | Upper | Lower |
| Overall, no covariates | | 0.0836 | 0.0717 | 0.0973 |
| Social gathering  0  1  0  1 | **Shared sanitary facilities**  **0**  **0**  **1**  **1** | 0.0209  0.0537  0.0565  0.1376 | 0.0105  0.0318  0.0341  0.1152 | 0.0412  0.0893  0.0923  0.1636 |

**Supplementary table 1: Infection probabilities with confidence intervals and influence of covariates in residence units harbouring an** **index case by generalized estimated equation analysis**

Abbreviations: CI = confidence interval, Infection probability = secondary infection probability, Lower = lower limit of the 95% CI interval, Shared sanitary facilities: the index case sharing facilities with other residents, Social gathering= the occurrence of a social gathering in the student residence which was attended by the index case. Upper = upper limit of the 95% CI interval. 0 = Not present, 1 = Present.

**Supplementary table 2: Infection probabilities and influence of covariates in residences harbouring an index case by generalized linear mixed model analysis.**

Using the GMLL model, the average probability of secondary infection was estimated at 0.0855, close to the GEE result. The 95% confidence interval in this model corresponds to the estimated range of secondary attack rates which are to be expected in individual residence units. This was estimated between 0% and 47%. Of all covariates placed in the GMLL model, only occurrence of a social gathering attended by the index case remained a significant risk factor (p=0.0019). The shared use of sanitary facilities was borderline not significant in this model (p=0.0552). However, we decided to keep it in for coherence between all approaches. As presented in table 3, in residence units without the occurrence of a social gathering attended by the index case and without the shared use of sanitary facilities, the range of secondary attack rates in the individual residence units was estimated between 0% and 17%. In the residence units with both occurrence of a social gathering in the residence attended by the index case and the shared use of sanitary facilities, the range of secondary attack rates was estimated between 1% and 59%. The other two groups again take an interim position between these two

| Residence units with index case present | | Infection  Probability | 95% Probability range | |
| --- | --- | --- | --- | --- |
|  |  |  | Upper | Lower |
| Overall, no covariates | | 0.0855 | 0.0015 | 0.4774 |
| Social gathering  0  1  0  1 | **Shared sanitary facilities**  **0**  **0**  **1**  **1** | 0.0282  0.0733  0.0616  0.1440 | 0.0006  0.0020  0.0017  0.0062 | 0.1685  0.3897  0.3316  0.5917 |

The 95% probability range corresponds to the range of expected secondary attack rates in individual residence units Abbreviations: . Infection probability = secondary infection probability, Lower = Lower limit of the 95% probability range, Shared sanitary facilities: the index case sharing sanitary facilities with other residents, Social gathering= the occurrence of a social gathering in the student residence which was attended by the index case, Upper = upper limit of the 95% probability range. 0 = Not present. 1 = Present.

**Supplementary figure 1**

**Supplementary figure 1: Timing of outbreaks in student residences.**

The left columns in this figure represent the number of student residences with secondary COVID-19 transmission following the diagnosis of a first index case. The right columns represent the number of student residences in which there was no secondary transmission. Most outbreaks occurred during March 2021 (44% (73/165)), corresponding to the third wave of infections in Belgium

Number of outbreaks

| Months | Student residencies with transmission | Students residencies without transmission |
| --- | --- | --- |
| 10-2020 | 2 | 1 |
| 11-2020 | 0 | 4 |
| 12-2020 | 2 | 2 |
| 01-2021 | 3 | 1 |
| 02-2021 | 6 | 9 |
| 03-2021 | 29 | 45 |
| 04-2021 | 11 | 21 |
| 05-2021 | 6 | 26 |
| 06-2021 | 0 | 3 |
